# Stairway visual contrast enhancement to reduce fall-related events

**DOI:** 10.1101/2022.03.25.22271716

**Authors:** Sara A. Harper, Christopher Long, Samantha Corbridge, Tyson S. Barrett, Alex Braeger, Brevin J. Zollinger, Amy E. Hale, Chayston B. Brown, Kenneth Harrison, Shandon L. Poulsen, Travis Boman, Christopher J. Dakin

**Affiliations:** Department of Kinesiology and Health Science, Utah State University, Logan, UT, USA; Sorenson Legacy Foundation Center for Clinical Excellence, Utah State University, Logan, UT, USA; Department of Psychology, Utah State University, Logan, UT, USA

**Author notes:** Correspondence: Christopher J. Dakin.

**Keywords:** Slips and falls, Accidents, human error, Interventions, Experimental Design, Motor Behavior

## Abstract

Falls on stairs occur frequently and pose a significant health and financial risk. Laboratory research has found that fall frequency can be reduced through contrast enhancement of the stairs by applying vinyl striping to the first and last steps’ faces, and all the steps’ top edges. Here we sought to determine if such step contrast enhancement can reduce the probability of fall-related events such as loss of balance, slips, trips, and falls in public university staircases. Cameras were used to capture stair users’ ascent and descent on contrast enhanced (‘striped’) and control stairways. Observed age group, observed gender, traverse direction (ascent, descent), fall-related events, and walking speed (m/s) were recorded. Fall-related events were reduced for the striped stairway compared to the control stairway (odds ratio = 2.87, average marginal effect = 0.002, p = .023) when controlling for observed gender, age category, and traverse direction. These results suggest contrast enhancement of first and last steps’ face and all steps’ edges may reduce fall-related events in public settings. Adding contrast enhancement to public stairways is a simple and cost-effective way to reduce the loss of balance, slips, and trips that could lead to catastrophic falls on stairs as well as the health and financial burden associated with such falls.

**PRECIS:** Contrast enhancement of stairway features may be a simple and cost-effective way to reduce falls on stairs. We show that monochrome striping on the first and last steps’ faces and black stripes on steps’ top edges reduce the probability of fall-related events in public stairways.

## INTRODUCTION

Approximately 3,000 people injure themselves falling on stairs every year in the United States, resulting in annual fall-related medical costs of approximately $92 billion (Blazewick et al., 2018). Moreover, individuals who have experienced one such fall are prone to fall again thus increasing the likelihood of serious injury (Masud & Morris, 2001). Thus, it is crucial that fall prevention strategies be researched and implemented to reduce the financial and health burden associated with falls on stairways.

The likelihood of a fall depends on a variety of factors affecting balance that are accentuated by visual, hearing, and muscular strength deficits (Elliott et al., 2015) associated with aging. Visual deficits, in particular, increase the likelihood of a fall on stairs (Brundle et al., 2015; Simoneau, Cavanagh, Ulbrecht, Leibowitz, & Tyrrell, 1991) because vision provides information about the environment that is necessary to adjust foot placement and foot clearance (Buckley, Timmis, Scally, & Elliott, 2011; Graci, Elliott, & Buckley, 2010). Often interventions such as strength and balance training are undertaken to prevent falls. However, these interventions typically require a significant commitment from the individual (Mian et al., 2007) and take time to meaningfully reduce the likelihood of falls (El-Khoury et al., 2013). In contrast, an environmental intervention, such as improving the visibility of features of the stairs, avoids the financial, time, and physical burdens associated with training interventions. In addition, environmental interventions such as stairway visibility improvements are often easily implemented in a public setting (Foster, Hotchkiss, Buckley, & Elliott, 2014).

By improving the visibility of the stairs, stair users’ gait can be altered to reduce the likelihood of fall. Some results of these gait alterations include increased toe clearance, increased heel clearance, faster walking speed, or reduced kinematic variance (Cohen & Sloan, 2016; den Brinker et al., 2005; D. B. Elliott, Foster, Whitaker, Scally, & Buckley, 2015; David B. Elliott, Vale, Whitaker, & Buckley, 2009; Foster, Buckley, Whitaker, & Elliott, 2016; Foster et al., 2014; Foster, Whitaker, Scally, Buckley, & Elliott, 2015; Jacobs, 2016; Schofield, Curzon-Jones, & Hollands, 2017; Shaheen et al., 2018; Skervin et al., 2021; Thomas et al., 2021; Zietz, Johannsen, & Hollands, 2011). In fact, Elliott, Foster and colleagues have demonstrated that vertical, monochrome striping on the face of the bottom and top steps can increase vertical foot clearance 0.8 cm for young adults and up to 2.1 cm for older adults (D. B. Elliott et al., 2015; David B. Elliott et al., 2009; Foster et al., 2016; Foster et al., 2015; Skervin et al., 2021), with 1.0 cm, a 17.5% improvement, in toe clearance observed in older adults. Importantly, introducing the vertical monochrome striping intervention to both the top and bottom steps maximized the vertical toe clearance effect, compared to no vertical striping, striping only the bottom step, or striping only the top step (D. B. Elliott et al., 2015; Foster et al., 2015). In addition, the effect of the monochrome vertical striping is also maximized when the spatial frequency of the stripes is approximately 12 cycles or stripes per meter compared to four, eight, 16 and 20 cycles per step (D. B. Elliott et al., 2015; Foster et al., 2015; Skervin et al., 2021).

Heel clearance can also be increased [(3.2 ± 1.9 cm to 4.7 ± 1.4 cm; effect size 1.07 (3)] by adding high contrast visual strips along the top front edge of all the steps (Cohen & Sloan, 2016; den Brinker et al., 2005; D. B. Elliott et al., 2015; Foster et al., 2014; Thomas et al., 2021; Zietz et al., 2011). The addition of high contrast edging to each step has been shown to increase heel clearance during descent (<5 mm) and reduce the number of times the foot accidentally contacts steps’ edges (D. B. Elliott et al., 2015). Importantly, while these two environmental interventions are independently effective, their combined effect may be greater than each intervention on their own (Foster et al., 2015).

Implemented together, these visual interventions represent a promising way to reduce locomotor behaviors that increase the likelihood of a fall. However, since these observations largely derive from laboratory settings, the size of these environmental interventions’ effect in public environments remains unclear. Specifically, laboratory investigations lack the wide range of influences and distractions present in public environments and it is unclear how large an effect the changes in toe and heel clearance observed in the lab have on the probability of a fall on public stairs (Edwards, Dulai, & Rahman, 2019). Therefore, public validation is an essential next step in determining the efficacy of such a visual, environmental intervention on reducing the probability of a fall.

Here, we sought to determine if a) enhancing contrast of the steps’ edges using black strips, and b) enhancing the visibility of the first and last steps’ face by adding vertical striping would reduce the probability of fall-related events such as loss of balance, slips, trips, and falls in a public setting. We compared the probability of observing fall-related events between a stairway with the contrast enhancements and a stairway without the contrast enhancements. We hypothesized that the probability of fall-related events (e.g., loss of balance, slips, trips, and falls) would be reduced in the intervention stairway compared to the control stairway.

## MATERIALS AND METHODS

### Participants

To test the efficacy of the contrast enhancement intervention, users of the East and West stairways in Utah State University’s Health, Physical Education and Recreation (HPER) building were recorded on video. The sample consisted largely of individuals attending Lifetime Wellness or Kinesiology courses (e.g., dance, yoga, and laboratory). It is important to note that the study was planned prior to the COVID-19 pandemic, when normally the HPER building has extensive programming, open to community members, including water aerobics, employee wellness aerobic and weight room, racquetball courts, and gymnasiums. Because data collection took place between September 2020 and February 2021, during the COVID-19 pandemic, the sample was skewed more toward university students than would usually be the case. Stair users appearing under the age of 18 were excluded from data collection (n = 2, See supplemental table) and the subsequent analysis. The study was approved by the institutional review board at Utah State University (Protocol #10773).

### Protocol

Four security cameras were placed in both the East and West stairways of the HPER building to capture stair users’ ascent and descent of the respective stairways. Security cameras (Lorex cameras, Lorex Technology Inc., Markham, CA, USA) were set to record based on motion detected prior to entry into the stairway and therefore video was captured for the entirety of the stair negotiation process. Signage was posted at the entry to both stairways indicating that security cameras were recording activity in the stairway. Video was recorded on a digital video recorder and recordings were transferred multiple times per week, via a secure universal serial bus (USB) device, to a secure cloud storage unit (Box, Inc. Redwood City, CA, USA). The East stairway consisted of 11 steps (0.16 – 0.17 m step height, 0.32 – 0.33 m in tread depth, with an overall hypotenuse length of 3.75 – 3.77 m) and the West stairway consisted of 13 steps (0.16 – 0.17 m step height, 0.32 – 0.33 m tread depth, and 4.49 – 4.53 m hypotenuse length).

### Intervention

Monochrome striping of the stair face consisted of 19 black vertical vinyl stripes (12 cycles/1 meter) on a white background along the face of the bottom and top stairs. Contrast enhancement of each steps’ edge consisted of 5.5 cm black vinyl strips (stair tread depth) applied to the top front edges of each step as done previously (D. B. Elliott et al., 2015; Foster et al., 2014; Foster et al., 2015). The control stairway was unaltered and used to compare to the contrast enhanced intervention stairway. After 5,000 individual stair users were coded, the stairway used as the intervention and control stairways were switched (counter-balanced) (**Figure 1**). The cost of the vinyl striping material was approximately $287.59 (USD) for the two stairways including counter-balancing.

**Figure 1.**
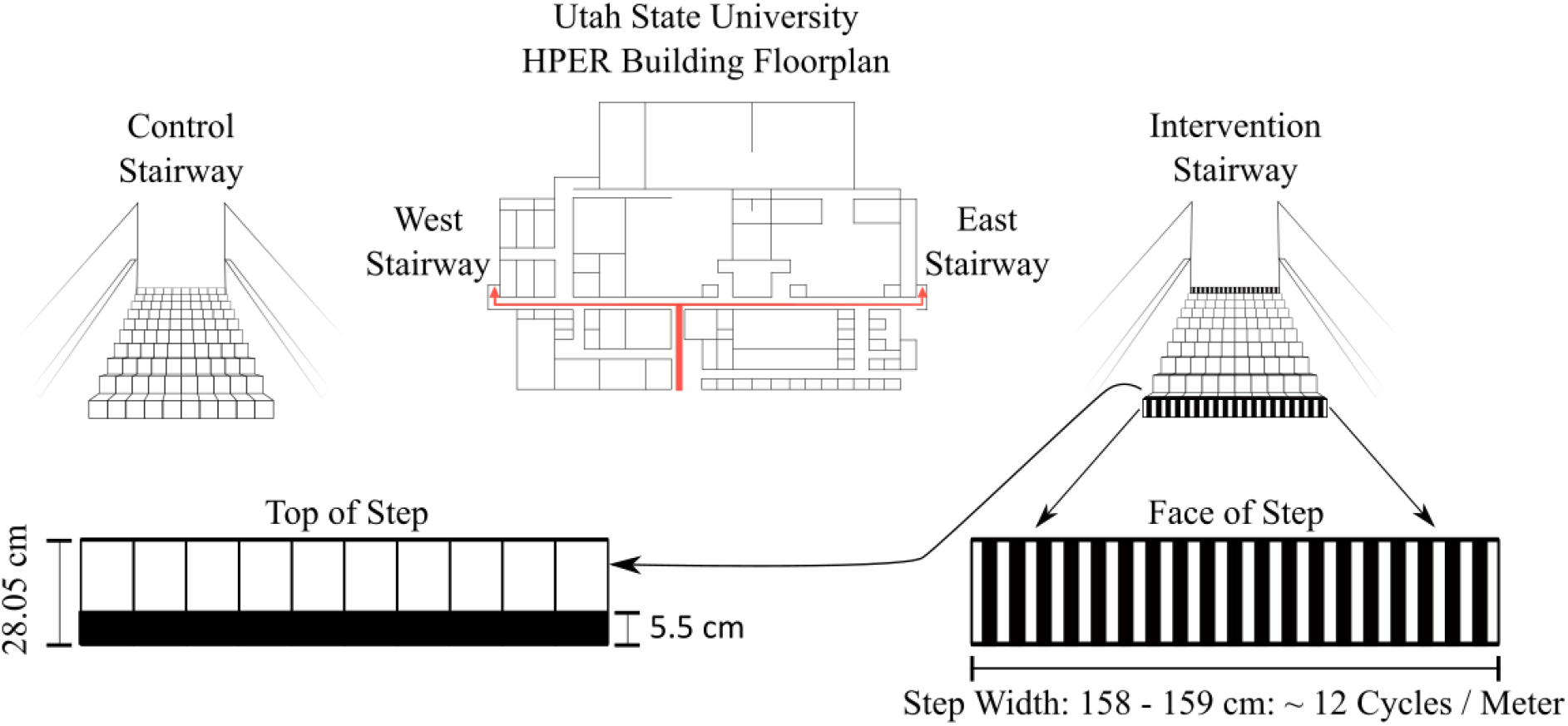
Study stairway locations. Schematic of the Health, Physical Education, and Recreation (HPER) building. Note: red arrows indicate the West and East stairways (upper middle). Visual representations of the control stairway condition (top left) and striping intervention (top right). Halfway through data collection, the intervention and control stairways were switched. The top front edge of steps in the intervention stairway were covered with a 5.5 cm black strip (bottom left) whereas the face of the first and last step of the intervention stairway were covered in a monochrome striping pattern with a spatial frequency of 12 stripes/meter (bottom right). The two stairways were approximately 112m apart.

In total, 12,285 individual observations were coded and balanced across each condition (intervention, control) and stairway (East, West). After accounting for incomplete observations, the complete data set included 2,500 individual observations coded for each of the conditions and stairways (i.e., East intervention, East control, West intervention, and West control each had 2,500 observations).

### Measures

Each outcome variable was assessed by stairway location (East stairway, West stairway) and condition (striping intervention, control). Both stairway light values were comparable between the upper and lower portions (approximately ∼19 lux average absolute difference). We coded stairway users based on their observed gender (men, women), estimated age range (18-40, 41-60, and 61+ years old), navigation direction (ascent, descent), date and time of day (24:00 clock), walking speed (m/s), and fall-related event severity (mild, moderate, and severe). A mild fall-related event represented a relatively subtle trip or slip with minimal recovery action (**Figure 2A**). A moderate fall-related event involved a loss in balance that required more substantial recovery such as a handrail reach (**Figure 2B)**. A severe fall-related event represented a complete loss in balance, resulting in a fall (**Figure 2C**). Figure 2 has been removed aligned with pre-print policy. We encourage readers to contact the corresponding author to request access to **Figure 2**.

Stair users, if traversing the stairs in a group, were documented as being in a group. To account for multiple repeat observations of stair users, we recorded time of day and date of each observation (further details below in Data Analysis). Data were recorded and encoded using Microsoft Excel (Microsoft Corporation, Redmond, WA, USA).

### Quality Analysis

To assess inter-coder reliability during data collection and encoding, each week one of the researchers would randomly select and evaluate ten percent of the data collected for that week. If an error was present, a second coder reviewed all data recorded by the first coder for the week in question and the second coder made a determination on the final coding record.

### Data Cleaning

Completed data entry consisted of 12,285 individual observations. In total, 1,148 observations were removed and are described in the supplemental information with 11,137 observations remaining for the final analysis. All observations were reviewed for missing or incomplete data. Next, observations were sorted by the time it took to traverse the stairs. All observations that took less than two seconds, or equal or greater than ten seconds to traverse the stairs were subject to additional review and removal to filter out unusual walking speeds. Next, individuals captured on video that were flagged for atypical events were reviewed and removed for the following reasons: use of assistive walking devices (e.g., walking boot, crutches, or knee brace), they were recorded outside of the designated data collection period (e.g., 17:01-6:59), or they did not use the stairs. In addition, all observations that involved stopping or pausing on the stairs or any unusual stair walking behaviors were removed (see Supplemental Information).

### Data Analysis

All descriptive statistics are presented as the number of observations and a percentage of the total observations. In order to assess differences by condition (intervention versus control), a series of generalized linear mixed effects models were used. Two models were tested to assess differences between the conditions. The first model assessed the main effect of condition on fall-related events. The remaining model tested for interactions between conditions. Covariates, such as gender, age category. All types of fall-related events were pooled, given their relative sparsity in the sample, and were coded as either present (a fall-related event occurred) or not. Given fall- related events were coded as binary, we used a Bernoulli (a special case of binomial) distribution and logit link for those models. For the fall-related events, these models can be expressed as:

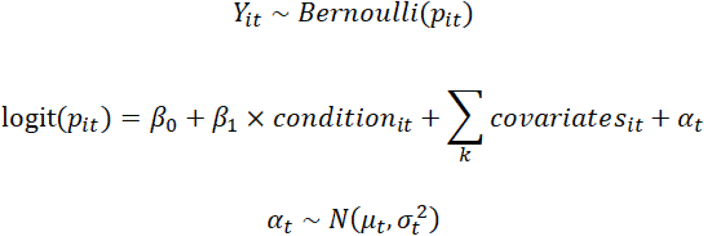

where *Y*_*it*_ is the binary outcome, *p*_*it*_ is the probability of a fall-related event occurring (i.e., Prob (*Y*_*it*_ = 1)) *²*_1_, is the estimate of interest, and *±*_1_ is the random intercept by hour of the day. The random intercept by hour of the day was selected to account for the clustering of individuals by certain time points as well as a proxy to help account for multiple observations of the same stair user given the challenge of identifying multiple observations of the same individual. Although imperfect, it explained variation in the outcome across models. Hypothesis significance testing for variables of interest was performed by comparing nested (i.e., more parsimonious) models using likelihood ratio tests (Hox, 2017). Estimates are highlighted in terms of both the odds ratio (relative differences) and the average marginal effect (absolute differences). Outcome measures from the model are presented using predicted probabilities. All statistical analyses were performed using R version 4.0.2. (Team, 2020) using the rio, tidyverse, janitor, lme4, lmerTest, furniture, and gtsummary packages (Barrett T, 2017; Chung-hong Chan, 2018; Daniel D. Sjoberg, 2020; Douglas Bates, 2015; Firke, 2020; Kuznetsova A, 2017; Wickham, 2019).

## RESULTS

Overall, most stair users appeared to be between the ages of 18-40 years (98.5%) with only 0.82% being between the ages of 41-60 and 0.68% appearing older than 61 years old. From this sample we observed a total of 20 fall-related events including 12 mild fall-related events, 4 moderate fall-related events, and 4 severe fall-related events. Of 11,137 complete observations, 12.82% (1,428/11,137) observations were verified twice. Across the sample, after accounting for gender, age category, and ascending/descending, there were differences in walking speed by age (p < .001), with observed young adults (18-40 years old) having the fastest stairway walking speed. Observed men also tended to walk faster than observed women (*p* < .001). The differences in walking speed did not depend on whether the individual was ascending or descending the stairway (*p* = .112).

**Table 1.** Participant characteristics.

| Outcome variables | Occurrences<br>(percentage of total) |
| --- | --- |
| Observed 18 - 40 years old | 10,970 (98.50 %) |
| Observed 41 - 60 years old | 91 (0.82%) |
| Observed 61+ years old | 76 (0.68%) |
| Observed women | 7,458 (66.97%) |
| Observed men | 3,679 (33.03%) |
| Mild fall-related event | 12 (0.108%) |
| Moderate fall-related event | 4 (0.036%) |
| Severe fall-related event | 4 (0.036%) |
| Total fall-related event | 20 (0.18%) |

Our primary aim was to examine whether the striping intervention reduced the probability of fall-related events. Likelihood ratio tests indicated there was a statistically significant difference (odds ratio = 2.87, average marginal effect = 0.002, *p* = .023), between the striped stairway (n = 6 fall-related events) and the control stairway (n = 14 fall-related events), with the striped stairway observing fewer fall-related events, on average, after controlling for observed gender, age category, and ascending/descending. None of the covariates were significantly related to fall-related events. The estimated differences by condition are presented in **Figure 3**, which shows the predicted probabilities of each condition experiencing a fall-related event. This difference did not depend on whether the individual was ascending or descending (*p* = .331).

**Figure 3.**
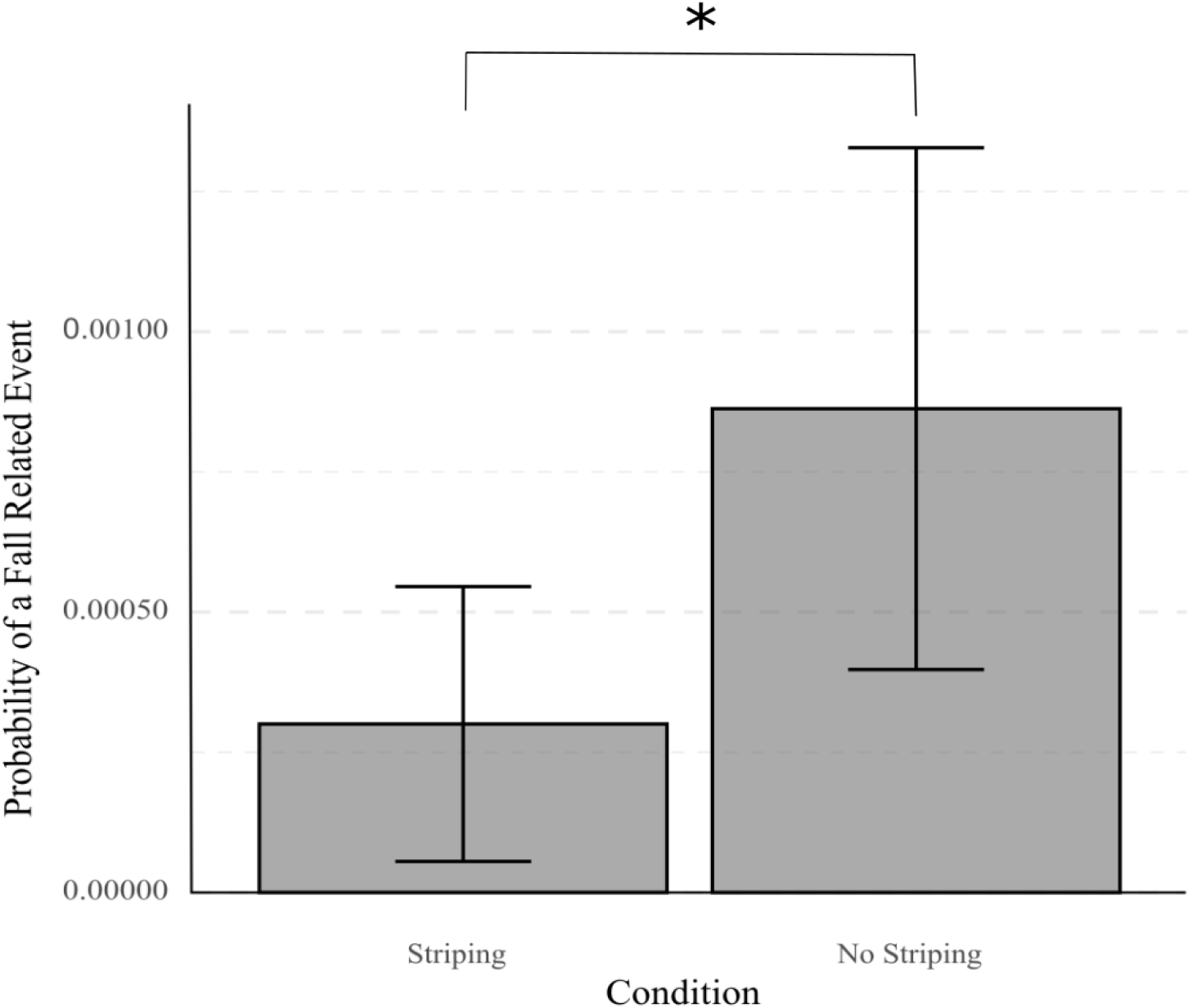
Stairway striping and fall-related events. Data are presented as estimated marginal mean ± 95% CI. Fall-related events decreased in the striped intervention stairways compared to the control stairways. * indicates a statistically significant difference as determined using likelihood ratio tests to compare the fit of the nested generalized linear mixed effects models *p* < .05.

## DISCUSSION

Here we sought to determine if contrast enhancement of features of the stairway can reduce the probability of experiencing a fall-related event in a public setting. We found that enhancing the edge and face contrast of stairs reduced fall risk factors compared to a control stairway without the intervention. These results suggest that this intervention may be a cost-effective way to reduce the loss of balance, slips, and trips that contribute to falls in public environments.

We found that the combined visual contrast enhancement of step face and edge reduced the number of fall-related events compared to a control stairway without the intervention. The fact that this effect was observed in our sample is notable since the sample was highly skewed toward a higher proportion of observed young adults (18-40 years old) due to the COVID-19 pandemic. While fall injuries are often reported among older adults, unintentional falls in young adults (18-35) represent the third most frequent causes of unintentional injury (Kool, Ameratunga, & Jackson, 2009). Young adults, in addition to older adults, could therefore also benefit from interventions meant to reduce fall-related events on stairways. Improvements in step contrast have been observed to increase foot step edge clearance in younger adults. In addition, younger adults with artificially induced visual impairments (20/200 vision) take smaller steps, and descend slower on low contrast stairways compared to high contrast stairways (David B. Elliott et al., 2009). Given that this intervention appears effective, at least to some degree, in younger adults, it is reasonable to believe it may be more effective in older adults, especially those with age-related declines in visual acuity (D. B. Elliott et al., 2015; Foster et al., 2014; Salvi, Akhtar, & Currie, 2006; Sjöstrand, Laatikainen, Hirvelä, Popovic, & Jonsson, 2011; Sjostrand, Bergman, & Popovic, 2004).

In a typical stairway, older adults with higher fall risk, determined by the Berg Balance Scale, Modified Falls Efficacy Scale, and Stair Self-Efficacy Questionnaire criteria, had both lower vertical tread clearance and lower step edge clearance compared to older adults with lower fall risk. This suggests that older adults at higher fall risk may benefit further from contrast enhancement than peers with lower fall risk. Interestingly, high visual contrast appears to not only increased foot clearance, but also reduced vertical acceleration of the center of mass in older adults with a higher likelihood of falling. Collectively, stairs with high visual contrast improved foot step edge clearance in both young and older adults, and also led to improved metrics of balance control in older adults with higher fall risk (Zietz et al., 2011).

One major benefit of recording stair users directly, compared to some prior fall assessment studies, is that the video recordings allow for a more objective assessment of fall-related events. With video, fall-related events can be assessed with greater resolution and without the subjective interpretation of the event from the individual that experienced the fall. In addition, through video review, smaller events that the individual experiencing the event might forget or omit can be identified. However, valuable information has also been derived from self-report of falls. For example, two studies have assessed falls in young adults using daily surveys that asked participants whether they had slipped, tripped or fallen. When the past 24 hours were queried (Cho, Heijnen, Craig, & Rietdyk, 2021; Heijnen & Rietdyk, 2016) 30% of students reported falling (excluding winter weather conditions) with 14% of those falls occurring on stairs (Heijnen & Rietdyk, 2016). In contrast when the prior 16-weeks were queried 52% of students reported falling with 13% occurring on stairs (Cho et al., 2021). Compared to national health surveys, 10.6% of adults ages 18-44 years old (Verma et al., 2016), and 18.5% of 20-45 years old (Talbot, Musiol, Witham, & Metter, 2005) reported falling over a one year period. The difference in percent of respondents reporting a fall between the daily surveys and the national health surveys may reflect the strengths of daily surveys, which may catch events forgotten or omitted from an annual report. In addition, our data suggest that falls may also be underreported. We observed that individuals fell once out of every five fall-related events or 20% of the time a fall related event occurred. In contrast, prior research indicates that students report falling, on average, once out of every 18 perceived slips or trips, a rate of 5.55% (Heijnen & Rietdyk, 2016). Similarly, a more recent investigation reported that 6% of slips and 4% of trips resulted in falls (Cho et al., 2021). Our observation appears in line with previous reports of discrepancies between video and survey data. Specifically, a study examining falls in long term care facilities found that video data and survey data tend to disagree about the cause of the fall 55.5% of the time (Yang, Feldman, Leung, Scott, & Robinovitch, 2015). This discrepancy demonstrates the value of incorporating video capture for documentation of fall-related events. While video recordings may improve objective slip, trip, and fall identification and classification, challenges using this medium still exist (Woolrych et al., 2015). Indeed, a considerable amount of personnel and time were required to code video recordings in our investigation, which could increase the cost of such video review compared to the use of surveys.

### Limitations

It is important to note that this study was planned prior to the COVID-19 pandemic, when normally the HPER building has extensive programming for all age groups and is open to community members (e.g., water aerobics, employee wellness aerobic and weight room, racquetball courts, and gymnasiums). However, data collection occurred during the COVID-19 pandemic, which reduced the number of mid-life and older adults that frequented the building leading to a sample skewed more towards young adults than was initially planned. This also likely led to fewer fall-related events. In addition, because our investigation observed stair users in a building that houses the Kinesiology and Health Science Department students, staff, and faculty, it is likely that the individuals we observed were more athletic on average, than non- users of this building. As a result of these limitations, it is possible that the effect size we observed is smaller than would be observed with a more heterogenous sample. Furthermore, because each stair user could not be directly queried, we estimated stair users’ age and gender in addition to introducing a date and time factor to account for multiple observations. It is important to note that because observed age and gender were estimated from video recordings, the precision of these measures is lower than if stair users were queried for these variables directly. Similarly, introducing a date and time variable is a less precise approach to account for multiple observations compared to having and using each individual’s identity. While our analysis did assess the time of day of repeat observations of stair users, time of day only explained approximately 1% of the variance in the data.

### Future directions

Although our investigation addressed our initial research question, additional questions now arise. Future work should consider observing a more heterogenous sample with a broader distribution of age ranges, emphasizing middle-age and older-age adult inclusion. Since falls disproportionally affect older adults, studies targeting older adults may provide additional insight into whether real life application of visual contrast enhancement of stairs will assist those most in need. In addition, future work could consider implementing machine learning fall detection software or programming which may eliminate the personnel resource for monitoring and detecting stair negotiation behavior and fall-related events (Shu & Shu, 2021).

## CONCLUSIONS

Here we built upon visual contrast stair inventions that have been evaluated in the laboratory by implementing such an intervention in a public setting (Edwards et al., 2019; D. B. Elliott et al., 2015; Jacobs, 2016; Skervin et al., 2021). We found that the enhancement of visual contrast of the first and last steps’ face and all steps’ edges reduced the number of fall-related events. Since the effect was still observed in a sample of primarily young adults, our results suggest that such visual contrast enhancements may represent a simple and inexpensive intervention to reduce loss of balance, slips, and trips that can lead to catastrophic falls on stairs across all populations. Such interventions could reduce the health and financial burden associated with falls on stairways.

## Data Availability

The data is not currently available.

## Acknowledgements

We would like thank Emmalee Rolfe, McKay Wilding, and Erika Larson for their assistance coding video observations and Utah State University Facilities for installing the video cameras and striping intervention.

## Supplemental Information

| **Removed observations (n=1,148)** | **Frequency (percentage)** |
| --- | --- |
| Outside of time frame (17:01-6:59) | 184 (16.03%) |
| Wrong camera angle (e.g., platform camera angle) | 120 (10.45%) |
| Partial device use (e.g., phone, smart watch) | 101 (8.80%) |
| Researcher in video (retrieving data files) | 80 (6.97%) |
| Missing stair start time | 71 (6.18%) |
| Reserve Officers' Training Corps on stairs | 66 (5.75%) |
| Assistant walking devices<br>(e.g., walking boot, crutches, knee brace, or limping) | 62 (5.40%) |
| Looked or waved at a camera | 48 (4.18%) |
| No stair use (did not step on stairs) | 45 (3.92%) |
| Stopped or paused on stairs | 45 (3.92%) |
| No individual in video (e.g., lighting change) | 36 (3.14%) |
| Maintenance (e.g., cleaning, janitor) | 21 (1.83%) |
| Jumped a portion of stairs | 18 (1.57%) |
| Partial stair use or partial video recording of stair use | 17 (1.48%) |
| Vending machine use | 11 (0.96%) |
| Pretending to fall or have balance issues | 7 (0.61%) |
| Stretched on stairs, sat on stairs | 6 (0.52%) |
| Played tennis on stairs, bouncing paddle ball | 4 (0.35%) |
| Reviewed stair striping | 4 (0.35%) |
| Unusual stair behavior | 3 (0.26%) |
| Slide down handrail | 3 (0.26%) |
| Takes shoes off on stairs or in socks | 3 (0.26%) |
| Dropped something on stairs | 3 (0.26%) |
| Talking to someone not on stairs | 2 (0.17%) |
| Observed age under 18 years old | 2 (0.17%) |
| Unconfirmed phone use | 2 (0.17%) |
| Switches phone use hand on stairs | 1 (0.09%) |
| Video and description do not match | 1 (0.09%) |

Note. Duplicate codes were present.

## KEY POINTS

- Contrast enhancement of features of stairways may be an effective way to reduce falls on stairs.
- We show that monochrome striping on the first and last steps’ faces and black stripes on steps’ edges reduced the probability of fall-related events in public stairways.

## Notes

The research support was provided by Utah State University Undergraduate Research & Creative Opportunity (URCO) Award (2020 – Christopher Long, Samantha Corbridge), American Heart Association (Postdoctoral Fellowship 20POST34990005 Sara A. Harper).

### Competing Interest Statement

The authors have declared no competing interest.

### Funding Statement

This study was funded by Utah State University Undergraduate Research & Creative Opportunity (URCO) Award (2020 Christopher Long, Samantha Corbridge), American Heart Association (Postdoctoral Fellowship 20POST34990005 Sara A. Harper).

### Author Declarations

The institutional review board at Utah State University gave ethical approval of this work (Protocol #10773).

